# Mathematical modelling of COVID-19 vaccination strategies in Kyrgyzstan

**DOI:** 10.1101/2021.12.21.21268200

**Authors:** Ainura Moldokmatova, Aizhan Dooronbekova, Chynarkul Zhumalieva, Aibek Mukambetov, Aisuluu Kubatova, Nurbolot Usenbaev, Ainura Kutmanova, Aman Osmonov, Shamil Ibragimov, Talant Abdyldaev, Luzia Freitas, Lisa J White

## Abstract

**Objectives:** In December 2020, an unprecedented vaccination programme to deal with the COVID-19 pandemic was initiated worldwide. However, the vaccine provision is currently insufficient for most countries to vaccinate their entire eligible population, so it is essential to develop the most efficient vaccination strategies. COVID-19 disease severity and mortality vary by age, therefore age-dependent vaccination strategies must be developed.

**Study design/Methods:** Here, we use an age-dependent SIERS (susceptible–infected–exposed–recovered–susceptible) deterministic model to compare four hypothetical age-dependent vaccination strategies and their potential impact on the COVID-19 epidemic in Kyrgyzstan.

**Results:** Over the short-term (until March 2022), a vaccination rollout strategy focussed on high-risk groups (aged >50 years) with some vaccination among high-incidence groups (aged 20–49 years) may decrease symptomatic cases and COVID-19-attributable deaths. However, there will be limited impact on the estimated overall number of COVID-19 cases with the relatively low coverage of high-incidence groups (15–25% based on current vaccine availability). Vaccination plus non-pharmaceutical interventions (NPIs), such as mask wearing and social distancing, will further decrease COVID-19 incidence and mortality and may have an indirect impact on all-cause mortality.

**Conclusions:** Our results and other evidence suggest that vaccination is most effective in flattening the epidemic curve and reducing mortality if supported by NPIs. In the short-term, focussing on high-risk groups may reduce the burden on the health system and result in fewer deaths. However, the herd effect from delaying another peak may only be achieved by greater vaccination coverage in high-incidence groups.

## Introduction

The pandemic caused by the emergence of the novel coronavirus disease 2019 (COVID-19) began in late 2019. The development of vaccines against COVID-19 may now change the course of the pandemic. However, it is unlikely that all countries will have immediate access to sufficient vaccine supplies due to the high demand worldwide that will initially exceed manufacturing capacities.

In 2020, Gavi, the Vaccine Alliance, in collaboration with the World Health Organization (WHO) and the Coalition for Epidemic Preparedness Innovations (CEPI) initiated COVAX, an international platform to ensure equitable access to COVID-19 vaccines globally and help countries mitigate the negative impacts of the pandemic on their national health systems and economies. This initiative will enable countries with limited resources to vaccinate approximately 20% of their population through the pooling of resources by participating countries and economies.(1)

At the time of writing this article, in addition to 226,000 doses of the Oxford–AstraZeneca vaccine provided by COVAX (2), the Kyrgyz Republic received approximately 1.5 million doses of the Sinopharm vaccine from China(3) and 80,000 doses of the Sputnik V vaccine from the Russian Federation.(4) This will cover about 30% of the adult population of the Kyrgyz Republic or around 19% of the total population. As of 9 August 2021, about 2.7% of the population in the Kyrgyz Republic was reported to be fully vaccinated and 5.6% to be partially vaccinated.(5)

Having a limited quantity of vaccine doses represents a dilemma for the government of the Kyrgyz Republic as to which population groups should be prioritized for the optimal protection of the entire population and to reduce the pressure on the national health system. Currently, the country is going through a third wave of the epidemic, with higher incidence rates compared with the two previous waves due to the emergence of new variants of SARS-CoV-2, the virus that causes COVID-19.(6) Although the death rate is lower than in the previous waves, the country’s health system is being overwhelmed by the large number of patients with COVID-19 requiring care. Thus, in July, nearly all hospitals treating patients with COVID-19 in the capital city, Bishkek, reported that both their general wards and intensive care units (ICUs) were at capacity.(7,8)

Current evidence suggests that COVID-19 disease severity and mortality vary by age(9–12); therefore, it is essential that any vaccination strategy takes this into account.(13) Some countries, mostly high income countries with high levels of COVID-19 vaccination coverage, have already shown the effectiveness of vaccination strategies with regards to reducing both morbidity and mortality.(5,14–18) Our study adds a Kyrgyz-specific perspective to the existing evidence on the potential effects of vaccination on the course of the COVID-19 pandemic in low- and middle-income countries (LMICs).(19–24) Specifically, we modelled four hypothetical age-dependent vaccination strategies to explore optimal options for reducing both mortality and healthcare demand in Kyrgyzstan. The model took into account the national health system capacity and vaccine availability in Kyrgyzstan.

## Methods

To conduct this modelling work, we used the web-based interface of a dynamic, age-structured SEIRS (susceptible–exposed–infectious–recovered–susceptible) model. The model was developed by the COVID-19 Modelling (CoMo) Consortium in collaboration with the Oxford Modelling for Global Health (OMGH) Group to examine the effect of various intervention packages on the course of the COVID-19 pandemic and related burdens on national health systems in more than 150 countries.(25,26) Details of the CoMo model framework, equations and parameters were reported by Aguas and colleagues.(27)

### Model parameters

The default model parameter values for describing the natural history of COVID-19 disease and the clinical course of infection were applied for Kyrgyzstan due to a lack of relevant country-specific evidence.(25) We also used the sixteen age groups defined in the CoMo model, with the probability of contacts within and between age groups based on the social contact matrices for 152 countries reported by Prem and colleagues.(28) For the demographic parameters, we used the 2019 United Nations World Population Prospects Report(29) and data from the Kyrgyz National Statistics Committee.(30) Data relating to daily new cases of COVID-19 and deaths were derived from the official online national COVID-19 resource.(31)

Hospital capacity parameters included the availability of surge and ICU beds and ventilators. We also used internal reports from the Kyrgyz Ministry of Health (MoH) on health system preparedness for the COVID-19 epidemic for the simulation. For the duration of hospitalized infection for each subcategory (surge, ICU and those receiving ventilation), we consulted with the Republican Hospital of Infectious Diseases, the National Hospital, and the Kyrgyz MoH.

### Baseline interventions

At the time of running the model, vaccination coverage in Kyrgyzstan had only reached about 2%; therefore, in the baseline scenario we only included non-pharmaceutical interventions (NPIs), which formed the bulk of responses to the COVID-19 epidemic in the country. These NPIs included self-isolation of symptomatic or laboratory-confirmed cases, case tracing, voluntary home quarantining of individuals who had come into contact with a COVID-19 case, social distancing, hand hygiene, mask wearing, school closure, working from home, travel bans, and lockdown. The timeline of interventions is shown in **Fig. 1**.

**Fig. 1.**
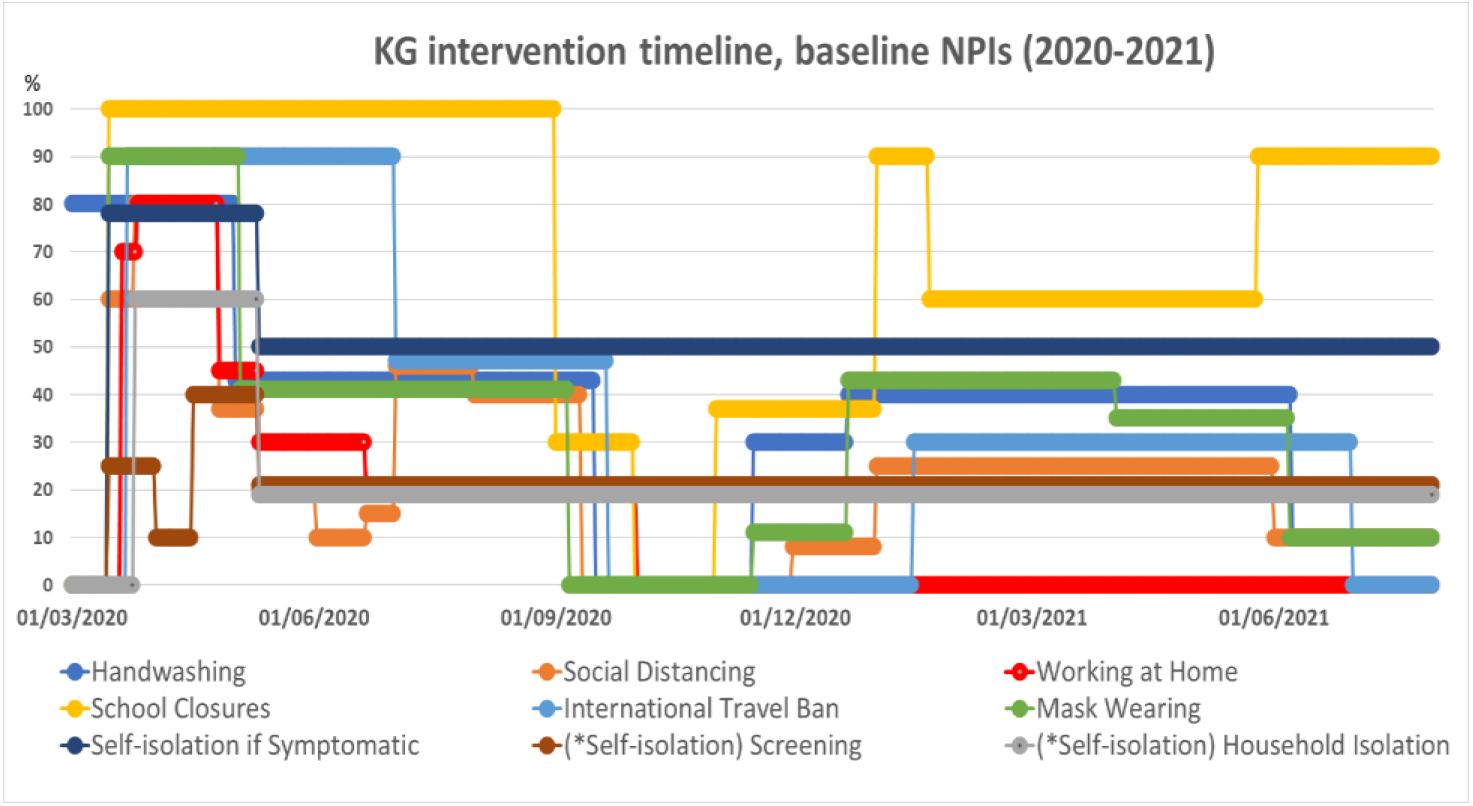
Timelines of the Kyrgyz government’s chosen NPI strategies and actual behavioural patterns (baseline scenario).

We defined the parameter values for Kyrgyzstan based on the estimated population behavioural patterns and prevention measures implemented in the country starting from the onset of the epidemic up to 15 June 2021. Reports and internal documents from the Kyrgyz Republic MoH, Ministry of Education, and Disaster Response Coordination Unit were used to estimate and define the values for school closures, travel bans, screening, household quarantining and self-isolation. As direct parameter values for social distancing and working from home were unavailable, we used data obtained from Google Community mobility reports for Kyrgyzstan(32) as a proxy. The data for mask wearing and hand hygiene were based on consultations with local health experts and sociologists.

The model assumes that if individuals with COVID-19 and their contacts self-isolated, then this would reduce transmission rates more than if these individuals did not self-isolate. It was also assumed that changes due to social distancing, as well as school closures and working from home, would reduce the level of social contacts within and between age groups, while the travel ban would reduce the number of imported cases.

### Hypothetical intervention scenarios

Four hypothetical scenarios for vaccination were explored in the simulation: 1) vaccination (65+ years, 70% coverage; 50–64 years, 40%; 20–49 years, 15%) and baseline NPIs; 2) vaccination (65+, 70%; 50–64, 40%; 20–49, 15%) and improved adherence to NPIs (mask wearing, 70%; social distancing, 25%); 3) improved vaccination coverage (65+, 70%; 50–64, 60%; 20–49, 25%) and baseline NPIs; 4) improved vaccination coverage (65+, 70%; 50–64, 60%; 20–49, 25%) and improved adherence to NPIs (mask wearing, 70%; social distancing, 25%). In each of these scenarios, we measured potential reductions in the numbers of new reported and unreported cases, deaths, and the burden on the health system, compared with these values in the baseline scenario. In addition, we estimated how NPIs could improve the effect resulting from the vaccination programme.

The vaccination coverage values are based on existing evidence for the age-related severity and mortality of COVID-19 disease, the planned supply of vaccines in the country(33) and the population structure, which is characterized by a comparatively smaller number of people of older ages.(29) After consulting with local experts, we also included changes of two NPIs in the model, i.e. adherence to mask wearing and social distancing, as these were considered to be affordable and less disruptive changes in terms of their effect on people’s economic and social lives. As shown in **Fig. 1**, mask wearing and social distancing are barely used in Kyrgyzstan.

It is important to note that the efficacy of mask wearing was assumed to be just 35% in the model, considering the comparatively low level of mask availability, their unaffordability, and people’s mask-wearing behaviour in Kyrgyzstan.

### Model validation

We validated the model validation is in two stages: 1) initial visual fitting and 2) particle filtering data fitting (PFDF). The fitting considered COVID-19 and various related factors, including hospitalization factors, intervention parameters, transmission probability given the number of contacts, the proportion of reported symptomatic and asymptomatic cases, reported hospitalizations, and the simulation start date. The details of the PFDF analysis and estimated parameter values can be found in the supplementary material.

The number of reported deaths is assumed to be more reliable than the number of confirmed new cases due to limited testing capacity and issues with the quality of tests in Kyrgyzstan; therefore, the fitting was carried out primarily against reported deaths, although we also performed the fitting against new cases. The predicted data calibrated against actual reported cumulative deaths and daily new cases are shown in **Fig. 2**.

**Fig. 2.**
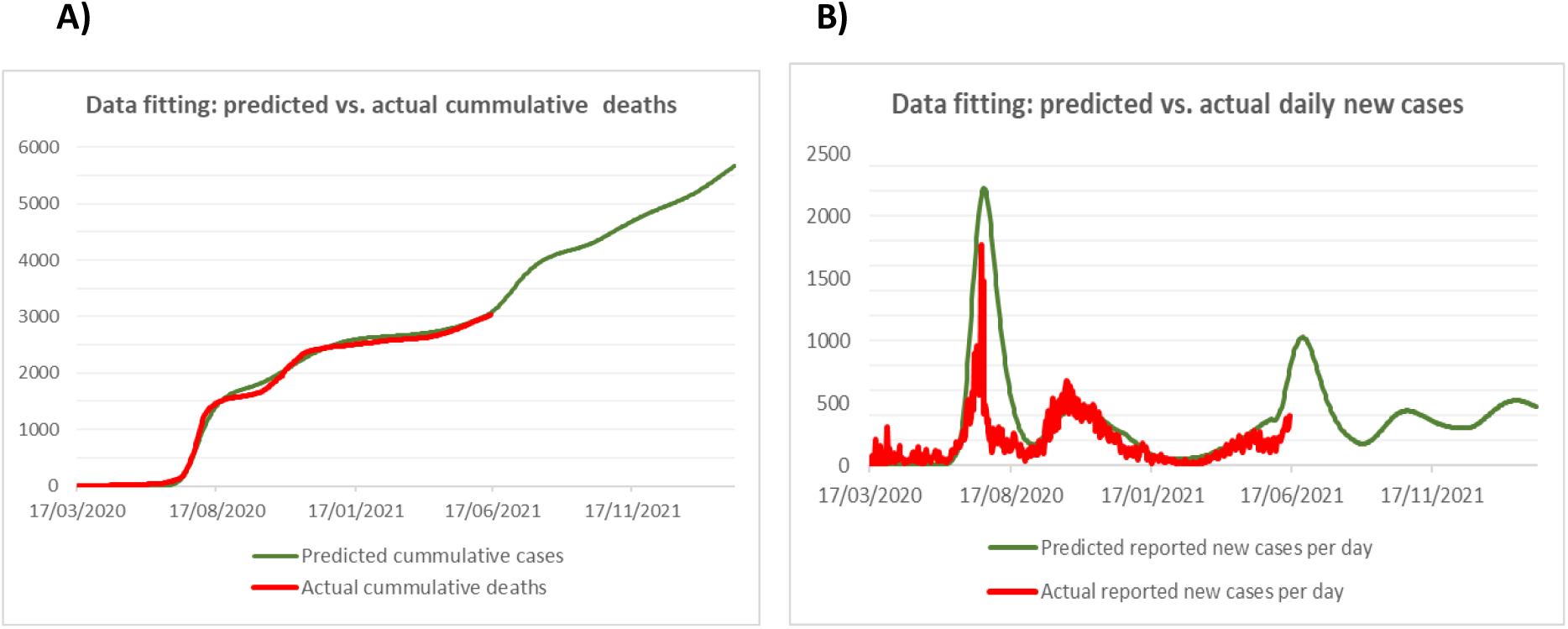
Particle filtering data fitting of the projected epidemic curve against actual reported cumulative deaths (A) and daily new cases of COVID-19 (B) as of 15 June 2021.

## Results

The results described here reflect the model’s projections of the impact of the baseline and the four hypothetical scenarios outlined above on the course of the COVID-19 epidemic, mortality, and the burden on the health system in Kyrgyzstan. In this modelling, we assumed that vaccination in all hypothetical scenarios began on 1 July 2021; therefore, the potential changes in the epidemic are projected starting from this date.

### New daily reported cases and estimated unreported cases

As shown in **Fig. 3B**, prioritizing the high-risk groups (i.e. those in the 50–64 and 65+ age groups) for the vaccination considerably decreases symptomatic and laboratory-confirmed cases of COVID-19 in all scenarios. Wider use of mask wearing and social distancing (scenarios 2 and 4) may accelerate the decrease in new cases in the current epidemic wave and flatten the peaks of any potential subsequent waves compared with alternative scenarios with baseline use of NPIs (scenarios 1 and 3).

**Fig. 3.**
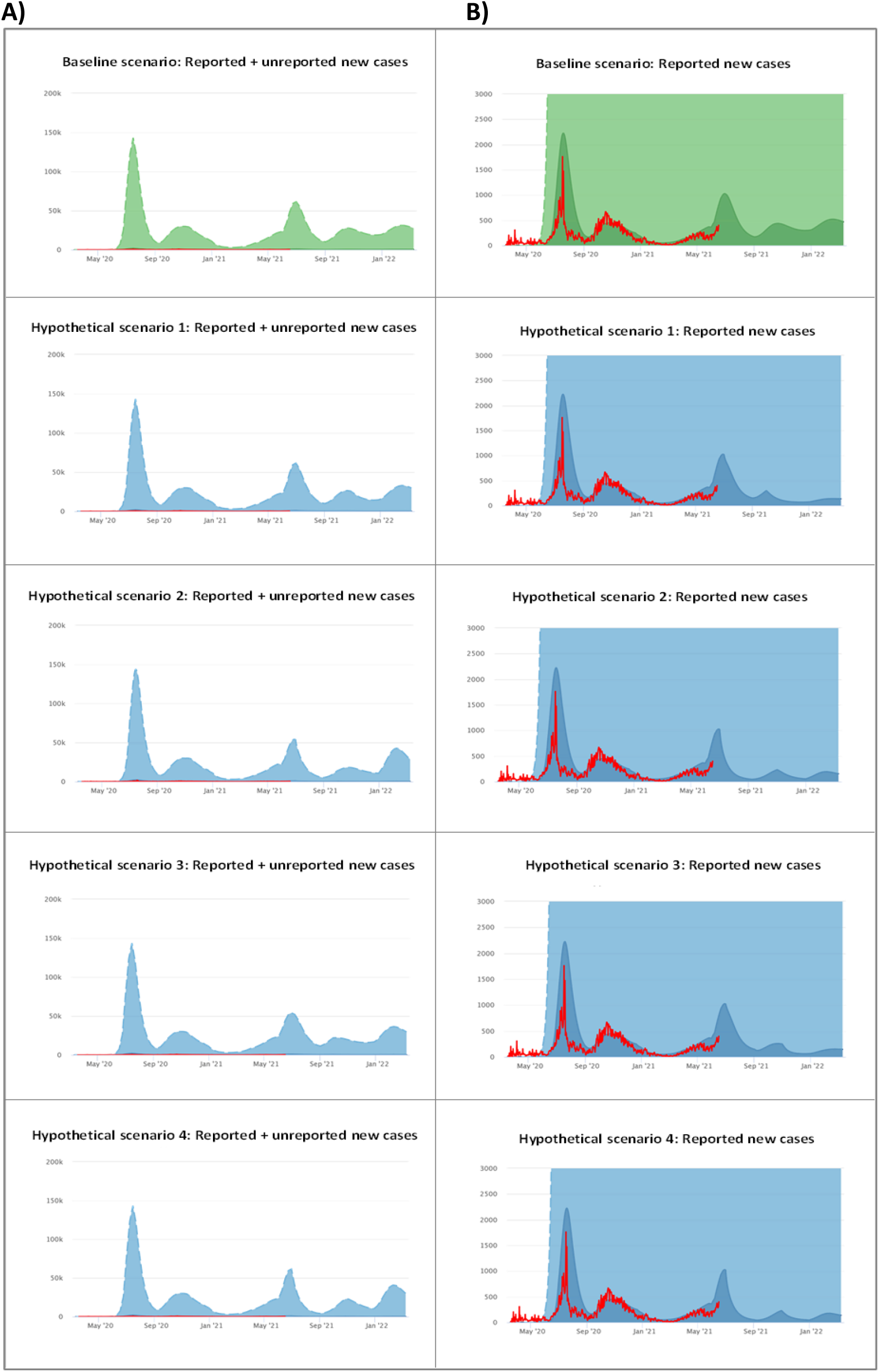
Baseline (A) and hypothetical scenarios (B) (1–4) of projected new daily reported and unreported cases of COVID-19 in Kyrgyzstan. The red curves represent the actual numbers of new daily reported cases.

The low vaccination coverage among high-incidence groups (aged 20–49 years), however, does not significantly change the number of unreported or asymptomatic cases in any of the scenarios (**Fig. 3A**), although the inclusion of improved use of two NPIs slightly reduces the peak of the next potential wave in October 2021, with an increase in the potential wave in February 2022, assuming that the indicated rates of vaccination coverage are completed by the end of the current year with no other rollout expected after that. In addition, as with reported cases, the improved use of NPIs may also accelerate the reduction in new unreported cases in the current wave.

### Burden on the health system

During the current wave of the COVID-19 epidemic, the occupancies of surge and ICU beds and ventilators in Kyrgyzstan may reach their capacity thresholds in both the baseline and hypothetical scenarios. However, the model predicts that the burden on hospitals will decrease considerably during the potential subsequent waves. Thus, according to Fig. 4, the occupancy of surge beds in the baseline scenario may reach between 1800 and 2100 during the subsequent waves, whereas in the hypothetical scenarios the occupancy may range between 300 and 1000.

**Fig. 4.**
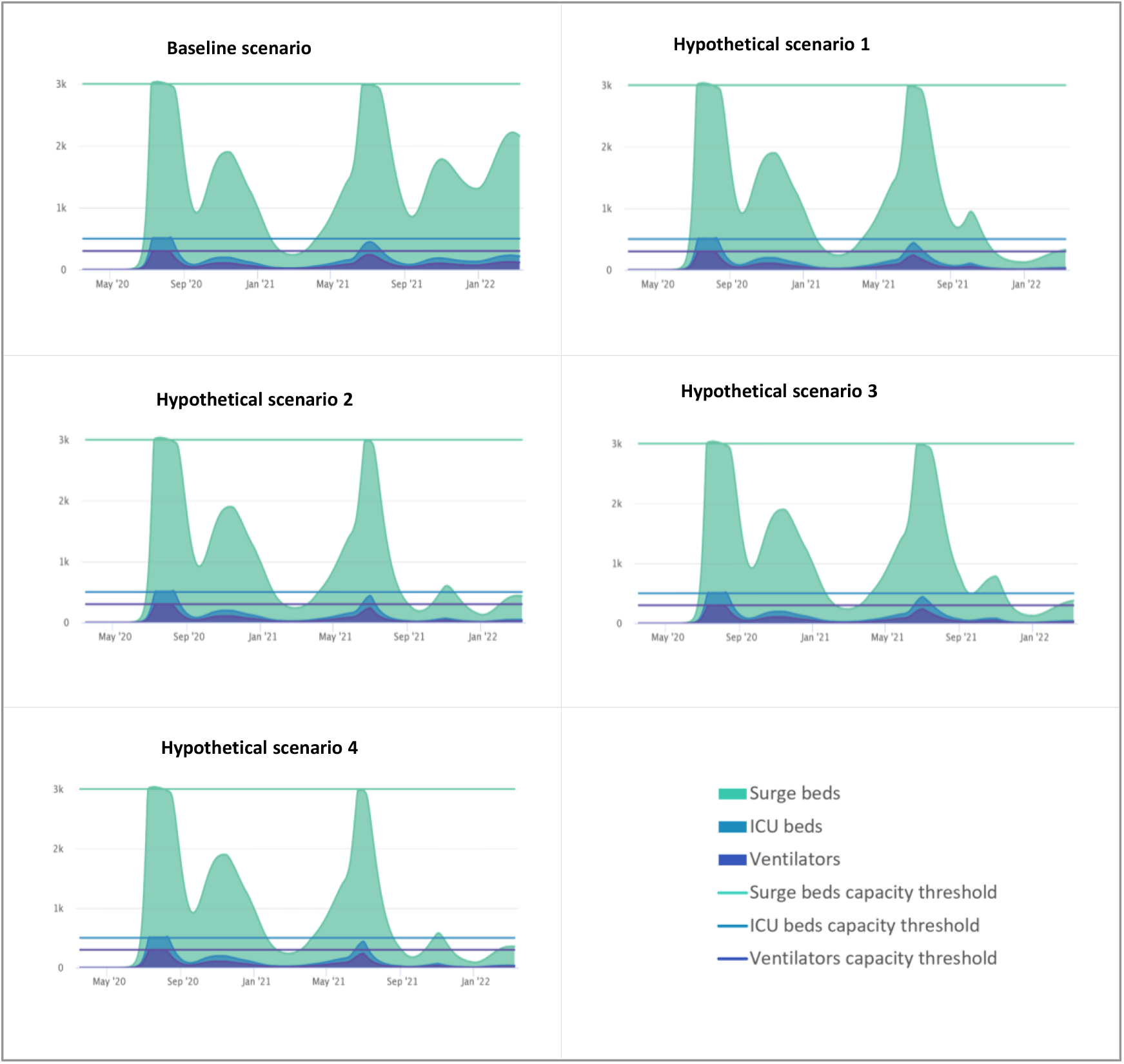
Baseline and hypothetical scenarios (1–4) of the projected burden of COVID-19 on the health system in Kyrgyzstan.

As with new cases, the greatest effect in decreasing the pressure on hospitals is likely to be achieved if vaccination is combined with improved mask wearing and social distancing. It is interesting to note that such an approach may also contribute to accelerating the reduction of the burden on the health system during the current wave (**Fig. 4**).

### Mortality

The model predicts that at the current level of NPIs and without vaccination (baseline scenario), the number of deaths attributable to COVID-19 will reach 7700 by the end of the model simulation period.

Vaccination and the resulting reduction in the pressure on the health system will decrease the number of COVID-19-attributable deaths. As shown in **Fig. 5**, the curve of the cumulative mortality of treated and untreated patients will flatten following the launch of the vaccination programme in all hypothetical scenarios.

**Fig. 5.**
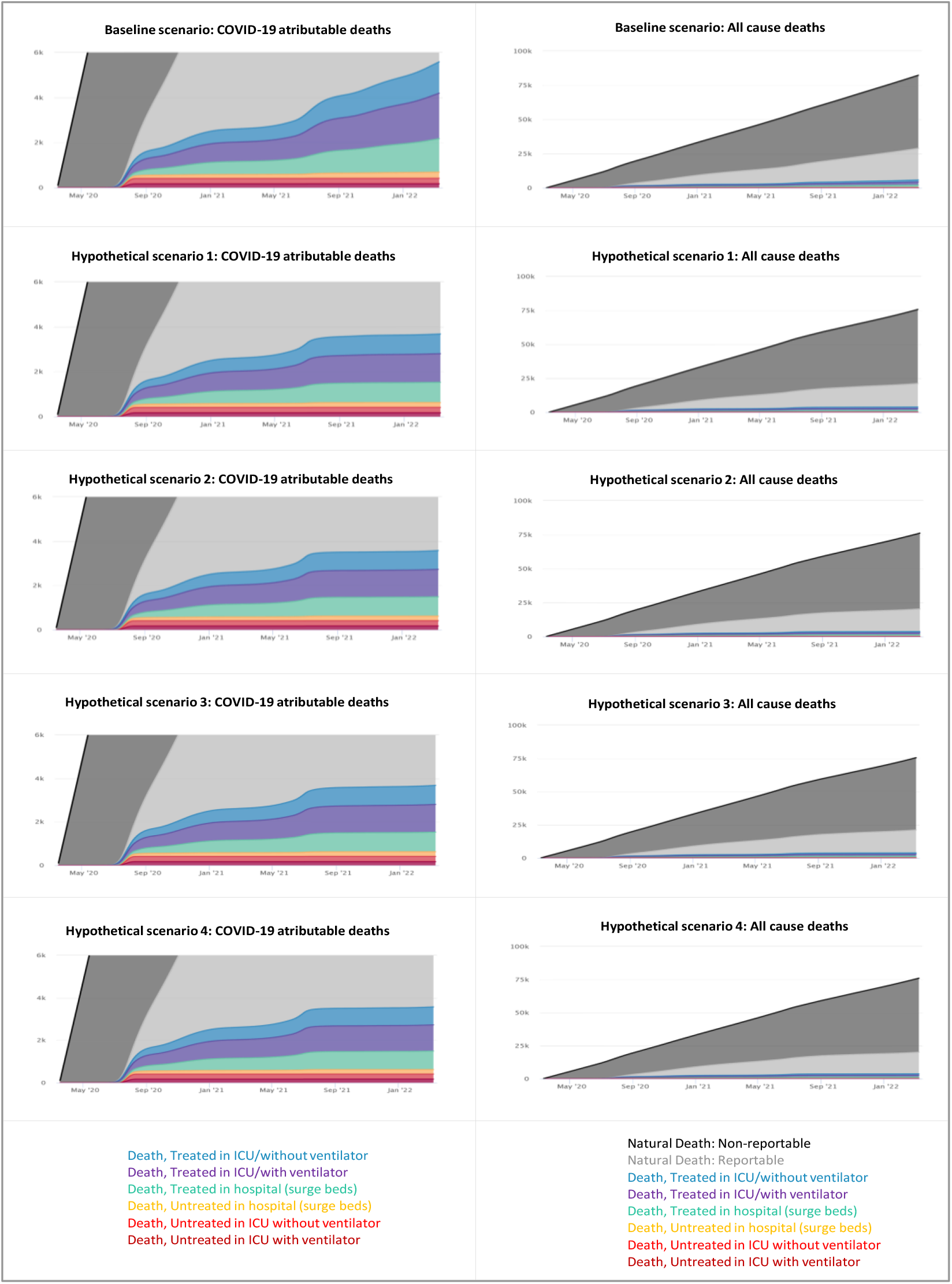
Baseline and hypothetical scenarios (1–4) of projected deaths attributable to COVID-19 and all-cause mortality.

The vaccination alone (scenarios 1 and 3) may reduce the number of deaths by 2040 and 2260 cases, respectively, while in combination with improved mask wearing and social distancing (scenarios 2 and 4), it may decrease mortality by 2480 and 2500 cases, respectively. In all scenarios, the majority of COVID-19-related deaths will occur in patients aged 65+ years (about 75%), followed by those aged 50 to 64 years (about 17%). This trend may be explained by the orgininally higher case fatality ratio (CFR) in these age groups, with a further increase during intensive phases of the epidemic **(Fig. 6)**.

**Fig. 6.**
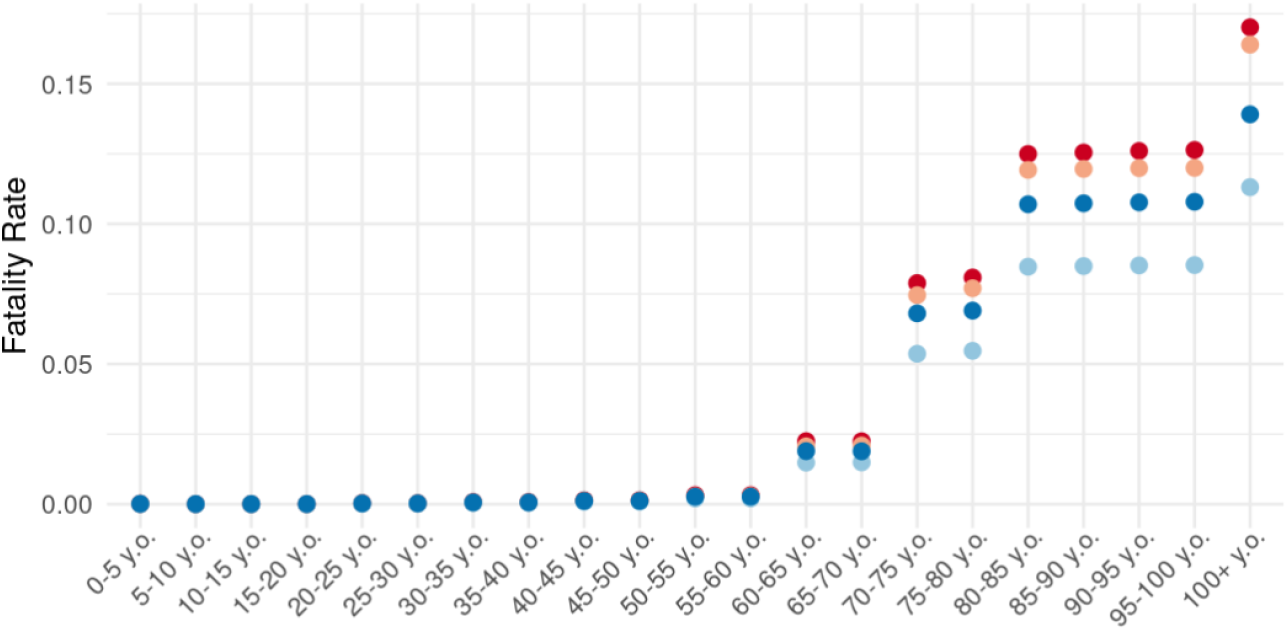
Changes in the case fatality ratio (CFR) during the course of the epidemic.

The vaccination programme, together with improved use of NPIs, may decrease all-cause mortality from 27,000 (baseline) to 16,000 cases (scenario 4), as hospitals and wards that had been reassigned to deal with COVID-19 patients will resume their work related to their original designation (**Fig. 5**).

## Discussion

The results obtained in this study suggest a potential positive impact over the short-term of age-specific vaccination in reducing the level of symptomatic cases of COVID-19, the burden on the health system, and mortality. According to the model’s outputs, prioritization of high-risk groups for vaccination may help to reduce COVID-19-attributable mortality and potentially have an indirect positive effect on reducing deaths from other causes as a result of the decreased burden on the health system.

It should be noted that the national health system in Kyrgyzstan, which has already deteriorated over the past 30 years, experienced tremendous pressure during the course of the COVID-19 epidemic. As in many other countries with limited resources, during the intensive phases of the epidemic, most of the available medical and human resources were reassigned and allocated to treat COVID-19 patients. As a result, the MoH reported 6430 (19.2%) cases of excess mortality in 2020, the majority of which were attributable to cardiovascular diseases, lung diseases and COVID-19.(34,35) Accordingly, the decreasing number of COVID-19 patients may help to improve the current situation in other areas of the health system.

On the other hand, considering that Kyrgyzstan has a young population, the primary focus of the vaccination programme on high-risk groups, with lower coverage among younger age groups, may have less effect in reducing the incidence level. Thus, the herd effect in delaying another peak may only be achieved by reaching high-incidence groups more broadly. Moreover, the model predicts that if vaccination is ceased once the indicated coverages are reached, this will trigger a later wave, equivalent to the baseline scenario, as the majority of the population will remain unprotected and the disease will continue to spread.

Finally, the model suggests that the combination of lower vaccination coverage with improved use of NPIs may have a similar effect to the combination of increased vaccination coverage with basic use of NPIs. In this model, we only considered increased coverage for mask wearing and social distancing, as these are considered to be less disruptive interventions than school closures or working from home. The model suggests that the combination of comparatively less disruptive interventions with vaccination may improve the effect of the vaccination programme. Considering that most countries with limited resources are not likely to receive sufficient supplies of vaccine in the near future,(1,36) such an approach could be of benefit for controlling the epidemic in these countries, at least in the short-term.

### Limitations

One of the major limitations of this work is that the model only accounted for the initial COVID-19 (“alpha”) variant, as at the time of the modelling the web-based application had no facility for projecting scenarios based on new variants of the virus. Accordingly, the actual effect on the epidemic and related health outcomes from a vaccination strategy may have different outcomes from that predicted depending on which variant(s) is prevalent in the country.

The other limitation is that due to the limited data available for the Kyrgyz context, some of the key parameter values we used for the simulations were based on assumptions and estimates. As mentioned earlier, in such cases we applied either existing global data, proxy national data or we estimated the values in consultation with local experts.

## Supporting information

Supplemental document: Model validation

## Data Availability

All data produced in the present work are contained in the manuscript

## Acknowledgement

We thank all who have collected, prepared, and shared data for this analysis. We are particularly thankful to our colleagues from the COVID-19 Modelling Consortium Dr. R.Aguas, Dr. O.Celhay and Dr. Bo Gao for the computer programming of the model and developing the web-based application, and Dr. Adam Bodley for the scientific editing of the manuscript.

