## Supplemental document: Model validation for "Mathematical modelling of COVID-19 vaccination strategies in Kyrgyzstan"

**Annex 1. Model validation**

**Part A. Particle filtering data fitting charts and tables**

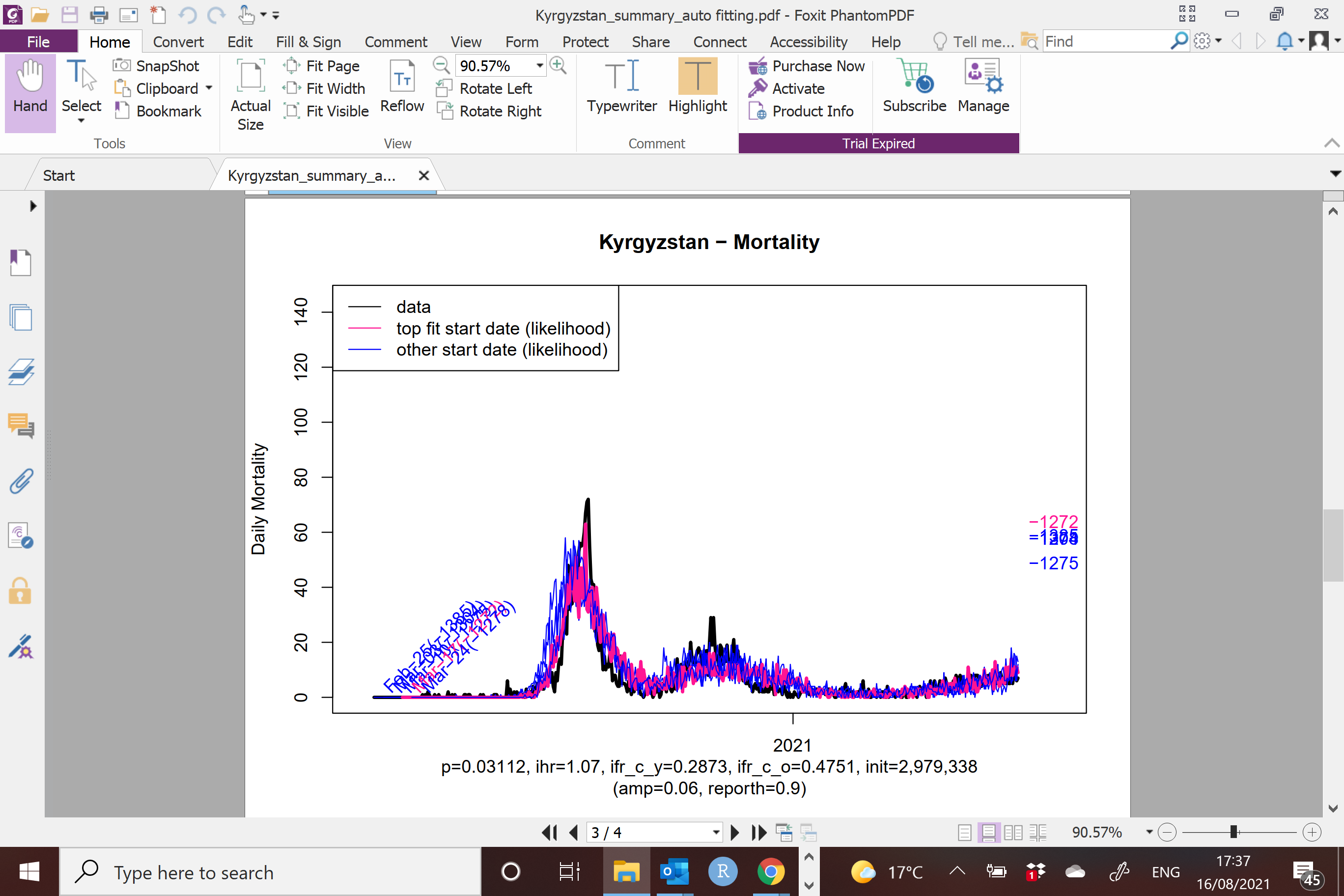

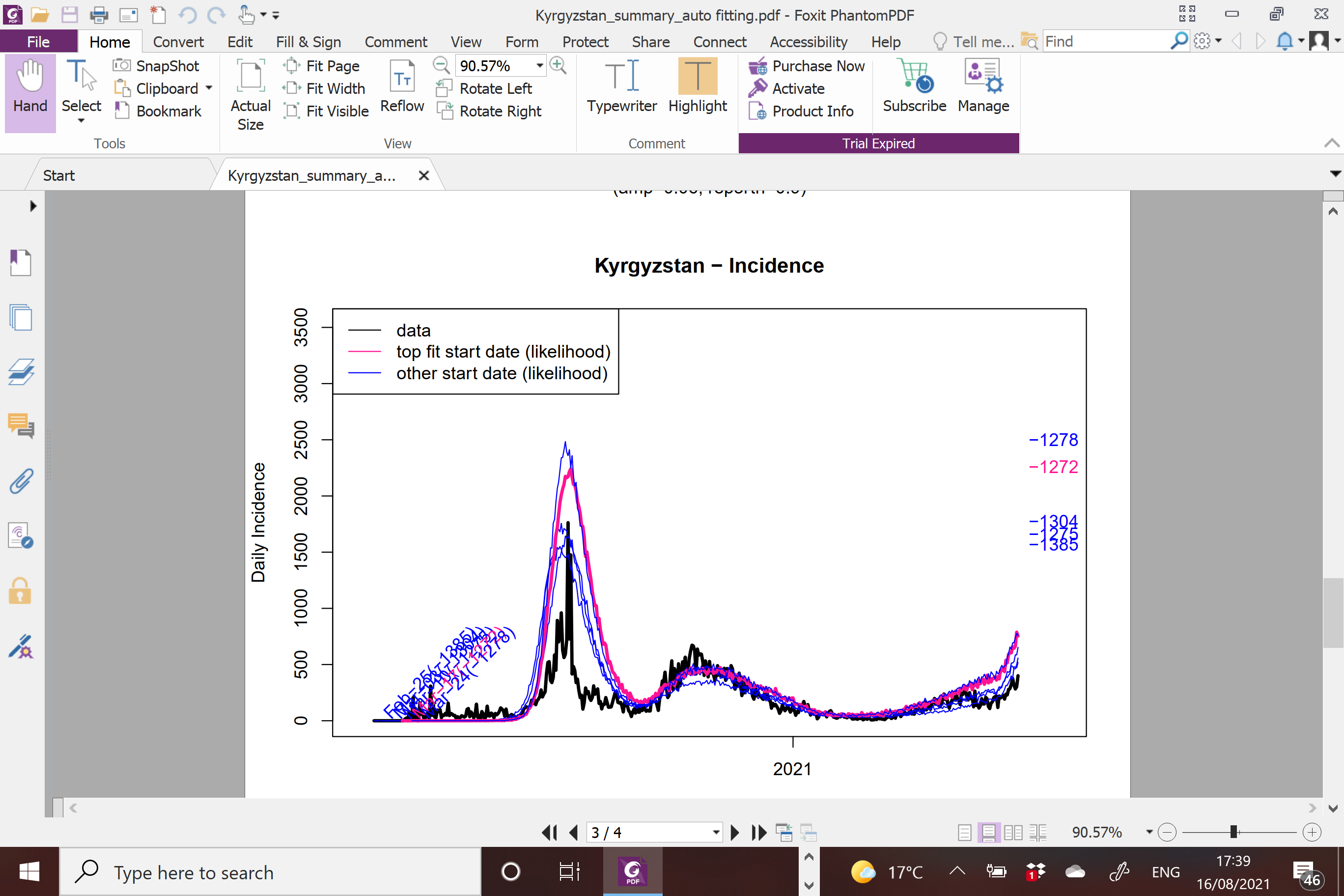

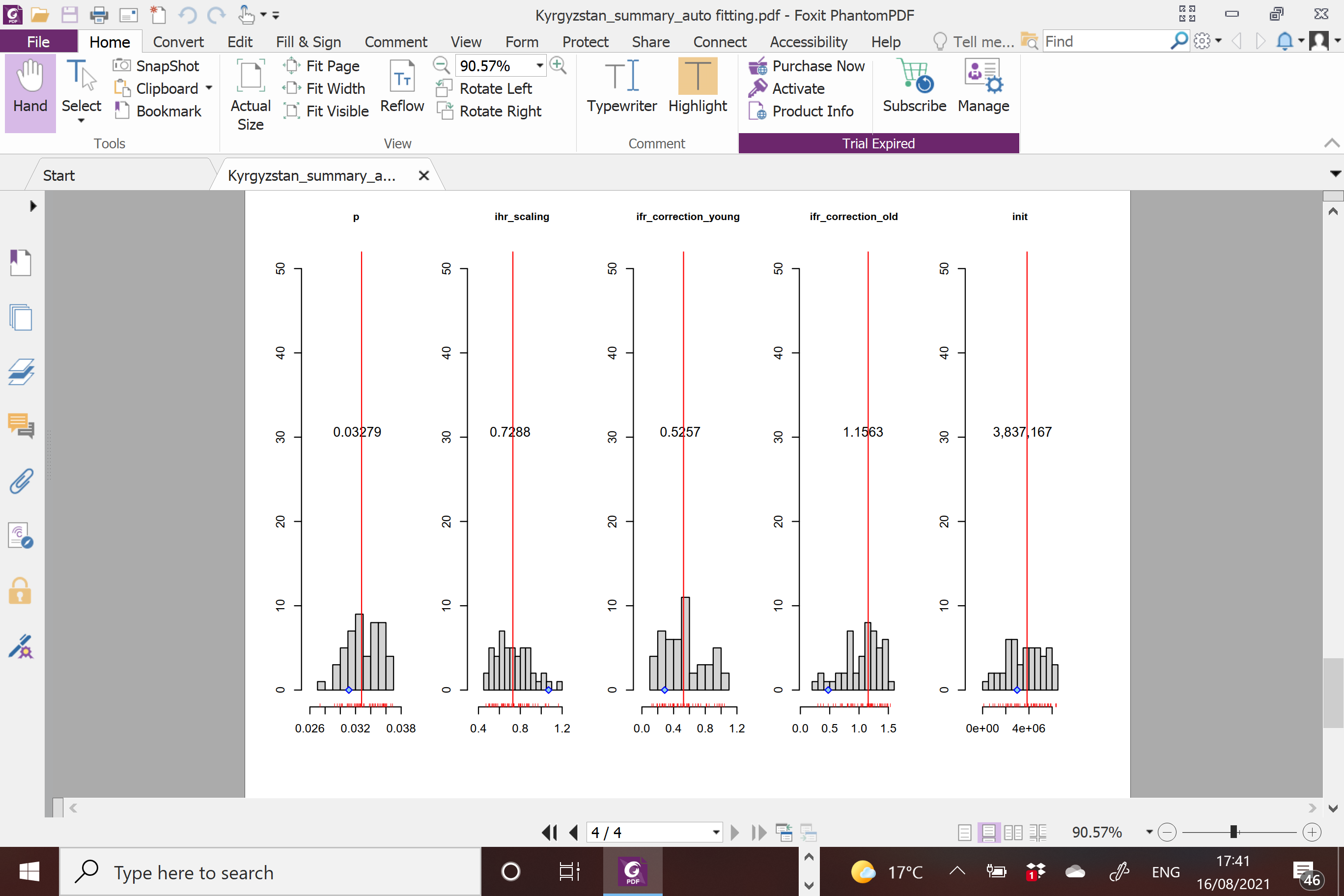

**Part B. Parameter values used in the particle filtering data fitting**

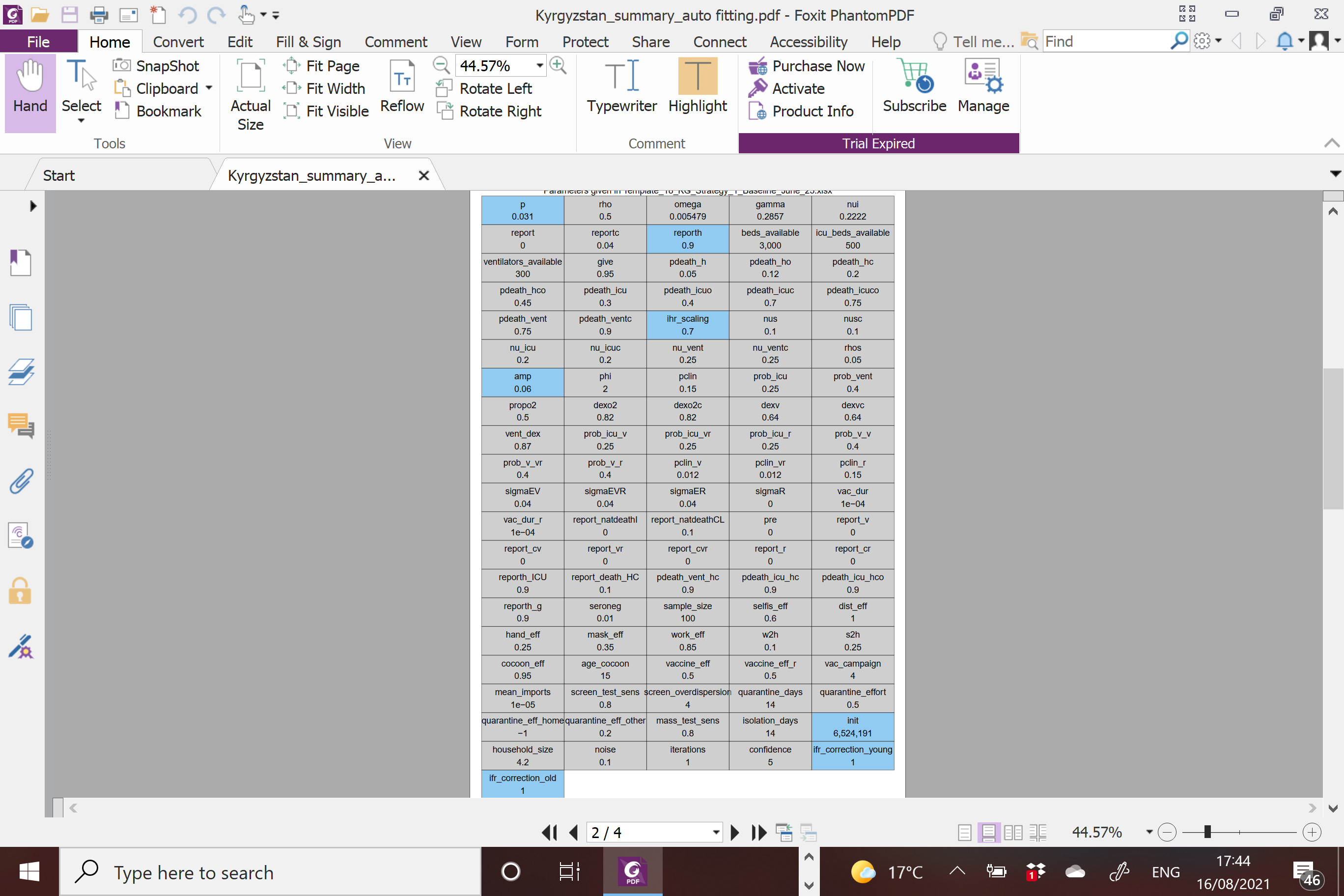

**Parameter code descriptions:**

| **Parameter description** | **Code** |
| --- | --- |
| Probability of infection given contact (0 to 0.2) | p |
| Relative infectiousness of incubation phase | rho |
| Average duration of immunity (0.5 to 150 days) | omega |
| Average incubation period (1 to 7 days) | gamma |
| Average duration of symptomatic infection period (1 to 7 days) | nui |
| Percentage of all asymptomatic infections that are reported | report |
| Percentage of all symptomatic infections that are reported | reportc |
| Percentage of non-severe hospitalizations that are appropriately treated | reporth |
| Maximum number of hospital surge beds | beds_available |
| Maximum number of ICU beds without ventilators | icu_beds_available |
| Maximum number of ICU beds with ventilators | ventilators_available |
| Probability of dying when hospitalized (not requiring O_2_) | pdeath_h |
| Probability of dying when hospitalized if requiring O_2_ | pdeath_ho |
| Probability of dying when denied hospitalization (not requiring O_2_) | pdeath_hc |
| Probability of dying when denied hospitalization if requiring O_2_ | pdeath_hco |
| Probability of dying when admitted to ICU (not requiring O_2_) | pdeath_icu |
| Probability of dying when admitted to ICU if requiring O_2_ | pdeath_icuo |
| Probability of dying when admission to ICU denied (not requiring O_2_) | pdeath_icuc |
| Probability of dying when admission to ICU denied if requiring O_2_ | pdeath_icuco |
| Probability of dying when ventilated | pdeath_vent |
| Probability of dying when ventilator denied | pdeath_ventc |
| Probability of dying when ventilator required and not going to hospital | pdeath_vent_hc |
| Probability of dying when ICU required (not requiring O_2_) and not going to hospital | pdeath_icu_hc |
| Probability of dying when ICU required (requiring O_2_) and not going to hospital | pdeath_icu_hco |
| Scaling factor for infection hospitalization rate: (0.1 to 5) | ihr_scaling |
| Duration of hospitalized infection: (1 to 30 days) | nus |
| Duration of ICU infection: (1 to 30 days) | nu_icu |
| Duration of ventilated infection: (1 to 30 days) | nu_vent |
| Relative percentage of regular daily contacts when hospitalized: | rhos |
| Month of peak infectivity of the virus (1, 2, …, 12) | phi |
| Annual variation in infectivity of the virus | amp |
| Probability upon infection of developing clinical symptoms | pclin |
| Probability upon infection of developing clinical symptoms if previously vaccinated | pclin_v |
| Probability upon infection of developing clinical symptoms if previously vaccinated and exposed | pclin_vr |
| Probability upon infection of developing clinical symptoms if previously infected | pclin_r |
| Probability upon hospitalization of requiring ICU admission | prob_icu |
| Probability upon admission to the ICU of requiring a ventilator | prob_vent |
| Proportion of hospitalized patients needing O_2_ | propo2 |
| Probability upon hospitalization of requiring ICU admission if previously vaccinated | prob_icu_v |
| Probability upon hospitalization of requiring ICU admission if previously vaccinated and exposed | prob_icu_vr |
| Probability upon hospitalization of requiring ICU admission if previously infected | prob_icu_r |
| Probability upon admission to the ICU of requiring a ventilator if previously vaccinated | prob_v_v |
| Probability upon admission to the ICU of requiring a ventilator if previously vaccinated and exposed | prob_v_vr |
| Probability upon admission to the ICU of requiring a ventilator if previously infected | prob_v_r |
| Relative risk of dying if needing O_2_ and taking Dexamethasone (Dex) | dexo2 |
| Relative risk of dying if needing ventilation and taking Dex | dexv |
| Relative risk of dying if needing but not receiving O_2_ and taking Dex | dexo2c |
| Relative risk of dying if needing but not receiving ventilation and taking Dex | dexvc |
| Change in ventilation requirement if given Dex | vent_dex |
| Probability of infection of people that have recovered from a previous infection | sigmaR |
| Probability of requiring hospitalization if previously vaccinated | sigmaEV |
| Probability of requiring hospitalization if previously infected | sigmaER |
| Probability of requiring hospitalization if previously infected and vaccinated | sigmaEVR |
| Vaccination - Duration of efficacious period | vac_dur |
| Vaccination - Duration of efficacious period if previously infected | vac_dur_r |
| Vaccination - Efficacy against infection | vaccine_eff |
| Vaccination - Efficacy against infection if previously infected | vaccine_eff_r |
| Vaccination - Time to reach target coverage (1 to 52) (weeks) | vac_campaign |
| Percentage of severe hospitalizations that are appropriately treated | reporth_ICU |
| Percentage of all asymptomatic infections in previously vaccinated people that are reported | report_v |
| Percentage of all asymptomatic infections in previously vaccinated and exposed people that are reported | report_vr |
| Percentage of all asymptomatic infections in previously infected people that are reported | report_r |
| Percentage of all symptomatic infections in previously vaccinated people that are reported | report_cv |
| Percentage of all symptomatic infections in previously vaccinated and exposed people that are reported | report_cvr |
| Percentage of all symptomatic infections in previously infected people that are reported | report_cr |
| Percentage of all people dying outside the hospital with asymptomatic infections reported as COVID-19-related deaths | report_natdeathI |
| Percentage of all people dying outside the hospital with symptomatic infections reported as COVID-19-related deaths | report_natdeathCL |
| Percentage of all people dying outside the hospital with severe infections reported as COVID-19-related deaths | report_death_HC |
| Days from seropositive to seronegative | seroneg |
| Average sample size for seroprevalence | Sample_size |
| Self-isolation if symptomatic: efficacy (0–1) | selfis_eff |
| Social distancing: efficacy (0–1) | dist_eff |
| Handwashing: efficacy: (0–0.25) | hand_eff |
| Mask wearing: efficacy (0–0.35) | mask_eff |
| Working from home: efficacy (0–1) | work_eff |
| Home contact inflation due to working from home | w2h |
| Home contact inflation due to school closure | s2h |
| Shielding the elderly: efficacy | cocoon_eff |
| Minimum age for elderly shielding: (0 to 100) | age_cocoon |
| Mass testing sensitivity | mass_test_sens |
| Mass testing: isolation days | isolation_days |
| Screen testing sensitivity | screen_test_sens |
| Screen testing overdispersion (1, 2, 3, 4 or 5) | screen_overdispersion |
| Household quarantine: days in isolation for an average person | quarantine_days |
| Household quarantine: days to implement maximum quarantine coverage: (1 to 5) | quarantine_effort |
| Household quarantine: decrease in the number of other contacts when quarantined: | quarantine_eff_other |
| Household quarantine: increase in the number of contacts at home when quarantined: | quarantine_eff_home |
| Number of exposed people at start date | init |
| Proportion of population with partial immunity at the start date | pre |
| Iterations (1 to 10,000) | iterations |
| Noise (0.01 to 0.2) | noise |
| Confidence (5 to 25) | confidence |
| Mean household size | household_size |
| Mean number of infectious immigrants per day | mean_imports |

**Details of age-distributed parameters**

A. **Population structure and birth and death rates**

| Age category (years) | Population | Number of births per person (i.e. 0.5* births per woman) per day | Deaths per person per day |
| --- | --- | --- | --- |
| 0–4 | 760,255 | 0.0 | 0.0000101605 |
| 5–9 | 769,195 | 0.0 | 0.0000007304 |
| 10–14 | 600,626 | 0.0 | 0.0000009053 |
| 15–19 | 500,075 | 0.0000448688 | 0.0000015779 |
| 20–24 | 514,389 | 0.0002696000 | 0.0000022355 |
| 25–29 | 568,551 | 0.0002296603 | 0.0000027381 |
| 30–34 | 572,187 | 0.0001459984 | 0.0000038920 |
| 35–39 | 442,518 | 0.0000888265 | 0.0000059593 |
| 40–44 | 361,459 | 0.0000250881 | 0.0000095105 |
| 45–49 | 324,976 | 0.0000012351 | 0.0000134190 |
| 50–54 | 295,973 | 0.0 | 0.0000201361 |
| 55–59 | 285,462 | 0.0 | 0.0000270330 |
| 60–64 | 220,069 | 0.0 | 0.0000408832 |
| 65–69 | 143,755 | 0.0 | 0.0000581985 |
| 70–74 | 78,619 | 0.0 | 0.0001209308 |
| 75–79 | 32,595 | 0.0 | 0.0004064232 |
| 80–84 | 32,507 | 0.0 | 0.0004353677 |
| 85–89 | 15,592 | 0.0 | 0.0004875172 |
| 90–94 | 4,435 | 0.0 | 0.0005742387 |
| 95–99 | 897 | 0.0 | 0.0006177715 |
| 100+ | 56 | 0.0 | 0.0098953750 |

B. **Age-based fatality ratio and infections that lead to hospitalization**

| Age category | Age-based relative fatality ratio in a well-resourced scenario (%) | Age-stratum-specific hospitalization (proportion of all (asymptomatic + symptomatic) infections that lead to hospitalization) (%) |
| --- | --- | --- |
| 0–4 | 0.0016 | 0 |
| 5–9 | 0.0016 | 0 |
| 10–14 | 0.007 | 0.04 |
| 15–19 | 0.007 | 0.04 |
| 20–24 | 0.031 | 1.1 |
| 25–29 | 0.031 | 1.1 |
| 30–34 | 0.26 | 3.43 |
| 35–39 | 0.26 | 3.43 |
| 40–44 | 0.48 | 4.25 |
| 45–49 | 0.48 | 4.25 |
| 50–54 | 0.6 | 8.2 |
| 55–59 | 0.6 | 8.2 |
| 60–64 | 1.9 | 11.8 |
| 65–69 | 1.9 | 11.8 |
| 70–74 | 4.3 | 16.6 |
| 75–79 | 4.3 | 16.6 |
| 80–84 | 7.8 | 18.4 |
| 85–89 | 7.8 | 18.4 |
| 90–94 | 7.8 | 18.4 |
| 95–99 | 7.8 | 18.4 |
| 100+ | 7.8 | 18.4 |
